# Persistent HBV replication and serological response during up to fifteen years of tenofovir-based antiretroviral therapy in HIV-hepatitis B coinfected patients: a multicenter prospective cohort study

**DOI:** 10.1101/2021.07.01.21259560

**Authors:** Lorenza N. C. Dezanet, Patrick Miailhes, Caroline Lascoux-Combe, Julie Chas, Sarah Maylin, Audrey Gabassi, Hayette Rougier, Constance Delaugerre, Karine Lacombe, Anders Boyd

## Abstract

**Objectives:** To determine the extent of hepatitis B virus (HBV) suppression and its association with hepatitis “e” antigen (HBeAg) and hepatitis B surface antigen (HBsAg)-seroclearance in HIV-HBV-coinfected patients undergoing long-term tenofovir (TDF)-based antiretroviral therapy (ART).

**Methods:** We prospectively followed 165 HIV-HBV-coinfected patients undergoing TDF-based ART. Serum HBV-DNA viral loads, HBeAg and HBsAg were obtained at TDF-initiation and every 6-12 months. We calculated the proportion achieving virological response (VR, <60 IU/mL) during follow-up. We also calculated rates of HBeAg- and HBsAg-seroclearance, which were compared between those who achieved versus never achieved VR during follow-up using an exact binomial test.

**Results:** During a median 8.1 years (IQR=4.0-13.2) of TDF-treatment, 152 (92.1%) patients were able to achieve VR and 13 (7.9%) never achieved VR (median HBV-DNA at the end of follow-up=608 IU/mL, range=67-52,400,000). The prevalence of individuals with detectable HBV-DNA (≥60 IU/mL) decreased during TDF-treatment: 15.1% (*n*=14/93) at 5-years, 3.2% (*n*=2/62) at 10-years and, 3.2% (*n*=1/31) at 15-years. 44/96 HBeAg-positive patients (6.15/100 person-years) had HBeAg-seroclearance and 13/165 patients overall (0.87/100 person-years) had HBsAg-seroclearance. No difference in HBeAg-seroclearance was observed between those who achieved versus never achieved VR (7.4 versus 3.7/100 person-years, *p*=0.33), while HBsAg-seroclearance was only observed in those with VR (1.0 versus 0/100 person-years, *p*=0.49; respectively). Individuals with VR also had a higher frequency of undetectable HIV-RNA during treatment (p<0.001).

**Conclusions:** During long-term TDF-based ART for HIV-HBV coinfection, persistent HBV viremia is apparent, but becomes less frequent over time. HBsAg-seroclearance only occurred in those with full HBV and relatively high HIV suppression.

## Introduction

In the past decade, liver-related mortality has continued to persist as one of the major causes of non-AIDS related deaths in HIV-positive patients.^1,2^ Coinfection with hepatitis B virus (HBV) has been implicated as a major reason for this finding.^3^ HBV infection by itself is associated with an increased risk of liver fibrosis progression, cirrhosis and hepatocellular carcinoma (HCC), which can be mitigated with effective HBV-DNA suppression.^4^ Given that tenofovir (TDF) has dual activity against HIV and HBV, long-term administration of TDF-containing antiretroviral therapy (ART) has been recommended for all HIV-HBV coinfected patients.^5,6^

Although TDF has a high genetic barrier to HBV resistance,^7^ at least 3 years of treatment may be required to achieve virological reponse,^8–10^ while 10-20% of HIV-HBV coinfected patients exhibit persistence of HBV replication during longer periods of TDF.^8,11–13^ Nevertheless, almost all studies to date evaluating HBV replication during TDF have followed patients for at most 5-10 years and consequently, it is uncertain what proportion of patients achieve suppression of HBV-DNA viral load with longer TDF-use.^8,14^

There are other therapeutic goals for improved prognosis, such as hepatitis B “e” antigen (HBeAg)-seroclearance (for those with HBeAg-positive serology) and importantly, hepatitis B surface antigen (HBsAg)-seroclearance. For TDF-treated HIV-HBV coinfected patients, almost half of those who are HBeAg-positive exhibit HBeAg-seroclearance and few overall attain HBsAg-seroclearance.^8,15,16^ Since HBeAg-seroclearance and HBsAg-seroclearance seem to only occur among HIV-HBV coinfected individuals undergoing TDF with sustained HBV virological response,^8^ there is concern regarding the consequences of persistent HBV replication on serological outcomes. Most of our understanding on HBV seroclearance rates also stems from studies of HIV-HBV coinfected and HBV-monoinfected patients with limited duration of TDF treatment.^8,14^

In this study, we aimed to evaluate the extent of HBV suppression and determinants of various forms of HBV persistence in patients coinfected with HIV-HBV undergoing up to 15 years of continuous TDF-based ART. We further intended to examine the relationship between HBV persistence and HBeAg-and HBsAg-seroclearance.

## Patients and Methods

### Study population

Patients were selected from the French HIV-HBV Cohort Study.^17^ Briefly, this longitudinal cohort study included 308 HIV-positive patients with chronic HBV infection from four centers located in Paris and Lyon, France. Patients were included if they had HIV-positive serological results confirmed by western blot and HBsAg-positive serological results for >6 months. Participants were recruited in 2002-2003 and followed up prospectively every 6-12 months until 2017-2018. The cohort design and procedures are described elsewhere.^17,18^

For this analysis, we included patients undergoing TDF-containing ART for ≥24 consecutive months. This timeframe was chosen since studies in HBV monoinfection and HIV-HBV coinfection have demonstrated that >90% achieve virological response within 24 months of nucleos(t)ide analogue (NA) therapy.^19^ We did not include patients with concomitant interferon/pegylated interferon (peg-IFN) or detectable hepatitis C virus (HCV) or hepatitis D virus (HDV) RNA.

### Ethics

All patients provided written informed consent to participate in the study and the protocol was approved by the appropriate ethics committee (Paris, France) in accordance with the Helsinki Declaration. ^17^

### Data collection

Demographic information was collected at study inclusion. HIV-related variables included HIV-RNA viral load (HIV-VL) and CD4^+^ cell count, and were collected before TDF-initiation and at each follow-up visit. HBV-related variables included HBV-DNA VL, alanine aminotransferase (ALT) levels, aspartate aminotransferase (AST) levels, qualitative HBeAg, anti-HBe antibodies, HBsAg, and anti-HBs antibodies, and were collected before TDF-initiation and at each follow-up visit. Cumulative exposure to viral replication was calculated using time-averaged copy-years over follow-up time (copy-years_TAVG_), as detailed elsewhere.^18^ At TDF-initiation, L-nucleoside-associated HBV mutations at positions rt173, rt180, and rt204 of the *pol* gene and at nucleotide 1896 of the *precore* gene were determined using DNA chip technology (bioMérieux, Marcy l’Etoile, France).^20^ Liver fibrosis was assessed at each yearly interval by the FibroTest® calculated from a standard battery of biochemical markers.^21^ METAVIR equivalents of this measure, as established in the HIV-HBV coinfected population, were used to grade liver fibrosis (F2=0.48-0.58, F3=0.59-0.73, F4≥0.74).^22^

### HBV replication profiles

HBV replication profiles were based on HBV-viral load (VL) at the end of the follow-up, as defined previously.^8^ First, patients were classified on whether or not they had undetectable HBV-VL at the last follow-up visit (HBV-DNA <60 IU/mL). Second, those with undetectable HBV-VL were divided into two subgroups: sustained virological response (sustained-VR; achieving and/or constantly maintaining HBV-DNA <60 IU/mL thereafter) or transient persistent viremia (PV; attaining <60 IU/mL, intermittently ≥60 IU/mL thereafter and returning to undetectable levels at the last visit). Patients with detectable HBV-VL at the last visit were divided into two subgroups: low-level persistent viremia (LL-PV; 60-2,000 IU/mL) or high-level persistent viremia (HL-PV; >2,000 IU/mL).

### Statistical analysis

Baseline was defined as the study visit at or directly before TDF-initiation. Follow-up began at TDF-initiation and continued until the last study visit, TDF-discontinuation, initiating peg-IFN, detection of HCV or HDV RNA, or death, whichever occurred first.

We used several sets of endpoints to evaluate HBV-DNA suppression. First, the percentage with undetectable HBV-DNA was calculated at the end of each yearly interval of follow-up. Second, the cumulative proportion achieving VR (HBV-DNA <60 IU/mL) was calculated during continuous time. Third, in the subset of patients who had achieved VR, we identified visits at which HBV-DNA VL was detectable (≥60/mL) after achieving VR [i.e. viral persistence]. Risk-factor analysis for viral persistence was performed on this subset of patients using follow-up that began at the first undetectable HBV-DNA VL and continued until right-censoring. Univariable odds ratios (OR) comparing the odds across levels of determinants over time and their 95% confidence intervals (95%CI) were calculated from a logistic regression model, which included a random-intercept to account for between-patient variation at baseline. A multivariable model was constructed by adding all covariables with a *p* value <0.20 in univariable analysis and removing nonsignificant variables in backward-stepwise fashion. Finally, we constructed HBV replication profiles as defined above. Comparisons between HBV replication profiles were performed for all clinical parameters at baseline and during follow-up using the Kruskal-Wallis test for continuous variables and Pearson’s χ^2^ test or Fisher’s exact test for categorical variables. Scatterplots and locally weighted scatterplot smoothing plots were used to illustrate the evolution of HBV-DNA replication according to HBV replication profiles.

We then used HBeAg-seroclearance (for HBeAg-positive patients) and HBsAg-seroclearance as endpoints. We estimated the cumulative proportion achieving these events, while only considering the first seroclearance event and not taking into account transitioning back to antigen-positive status. We estimated time to seroclearance using Kaplan-Meier curves and the incidence of seroclearance rates, which were compared between those who achieved versus never achieved VR using a two-sided Exact binomial test with mid-p assumption.

All statistical analyses were performed using STATA software (v15.1; College Station, Texas, USA) and significance was determined using a *p* value < 0.05.

## Results

### Description of the study population

Of the 308 patients included in the cohort, 143 were not included in analysis for the following reasons: did not initiate TDF (*n*=51), used TDF for less than 24 months (*n*=42), used concomitant PEG-IFN or IFN (*n*=20), ever had anti-HCV and/or anti-HDV antibody positive serology (*n*=23), or had insufficient information at baseline (*n*=7). Thus, 165 individuals were included in the present analysis (Supplementary Figure 1).

Of these 165 patients, most were male (83.6%) with a median age of 41.7 years (IQR=36.4-48.3) at TDF-initiation (Table 1). Almost all patients had previously initiated ART (99.4%), with a median 7.0 years of ART exposure (IQR=4.4-9.2), and thus median CD4+ count was 410/mm^3^ (IQR=288-596) and 97 (58.8%) had undetectable HIV-RNA. At baseline, 96 (58.2%) patients were HBeAg-positive and HBV-DNA was detectable in 75.2% of participants. Of the 147 patients (89.1%) with previous lamivudine (LAM) exposure, median LAM duration was 5.1 years (IQR=2.8-6.8) at TDF initiation and, among those with available information, 20/100 (20.0%) had baseline LAM resistance mutations. HBV genotype was determined in 105 patients, and most harbored genotype A (67.6%), followed by G (15.2%), E (8.6%) and D (8.6%). Among patients with available data on Fibrotest scores at TDF initiation (n=147), 36 (24.5%) had F3-F4 fibrosis.

**Table 1.** Baseline characteristics among HIV/hepatitis B virus cohort participants (N=165).

| Characteristic | N (%) or median (IQR) |
| --- | --- |
| Demographics |  |
| Gender, male/female (% male) | 138/25 (83.6) |
| Age, years* | 41.7 (36.4-48.3) |
| BMI $\geq 25$ Kg/m<sup>2</sup>, N=149* | 27 (18.1) |
| Born in high HBV endemic zone<sup>†</sup> | 45 (27.3) |
| HIV characteristics |  |
| Ever having an AIDS-defining illness<sup>†</sup> | 41 (24.9) |
| Known HIV infection duration, years* | 11.1 (7.10-15.0) |
| Detectable HIV-RNA<sup>†</sup> | 68 (41.2) |
| HIV-RNA<sup>‡</sup>, log<sub>10</sub> copies/mL* | 4.1 (2.9-4.6) |
| CD4<sup>+</sup> cell count, per mm<sup>3</sup>, N=164* | 410 (288-596) |
| CD4<sup>+</sup> nadir cell count, per mm<sup>3</sup>, N=152* | 224 (104-319) |
| Duration of prior ART, years, N=164* | 7.0 (4.4-9.2) |
| Viral hepatitis B characteristics |  |
| Known HBV infection duration, years, N=164* | 8.3 (4.2-12.1) |
| Prior LAM exposure<sup>†</sup> | 147 (89.1) |
| Cumulative prior LAM treatment, years, *N*=147* | 5.1 (2.8-6.8) |
| LAM-resistant mutations, *N*=100<sup>†</sup> | 20 (20.0) |
| Detectable HBV-DNA<sup>†</sup> | 124 (75.2) |
| HBV-DNA<sup>‡</sup>, log<sub>10</sub> IU/mL* | 5.2 (3.2-7.2) |
| HBeAg-positive<sup>†</sup> | 96 (58.2) |
| ALT, IU/L, *N*=161* | 40 (26-68) |
| AST, IU/L, *N*=161* | 34 (25-56) |
| F3-F4 fibrosis<sup>#</sup>, *N*=147<sup>†</sup> | 36 (24.5) |
| *Precore* mutations, *N*=101<sup>†</sup> | 22 (21.8) |
| HBV genotype, *N*=105<sup>†</sup> |  |
| A | 71 (67.6) |
| D | 9 (8.6) |
| E | 9 (8.6) |
| G | 16 (15.2) |

\*Median (IQR).
<sup>†</sup> Number (%).
<sup>#</sup> Estimated using the FibroTest®.
Abbreviations: AIDS, acquired immune deficiency syndrome; ALT, alanine aminotransferase; ART, antiretroviral therapy; AST, aspartate aminotransferase; BMI, body mass index; HBV, hepatitis B virus; HIV, human immunodeficiency virus; HBeAg, hepatitis B “e” antigen; LAM, lamivudine.

### HBV-DNA suppression during TDF-containing ART

Patients were followed for a median 8.1 years (IQR=4.0-13.2), with a maximum follow-up of 15.7 years. Three patients switched from tenofovir disoproxil fumarate to tenofovir alafenamide (TAF) based ART during follow-up, while follow-up during TAF was still included in analysis (median TAF duration: 0.5 years, range=0.2-5.6). The percentage of patients with undetectable HBV-DNA increased substantially in the first 6 years of TDF treatment, while, thereafter, ranging between 94.0-97.8% and never reaching 100% (Figure 1a).

**Figure 1.**
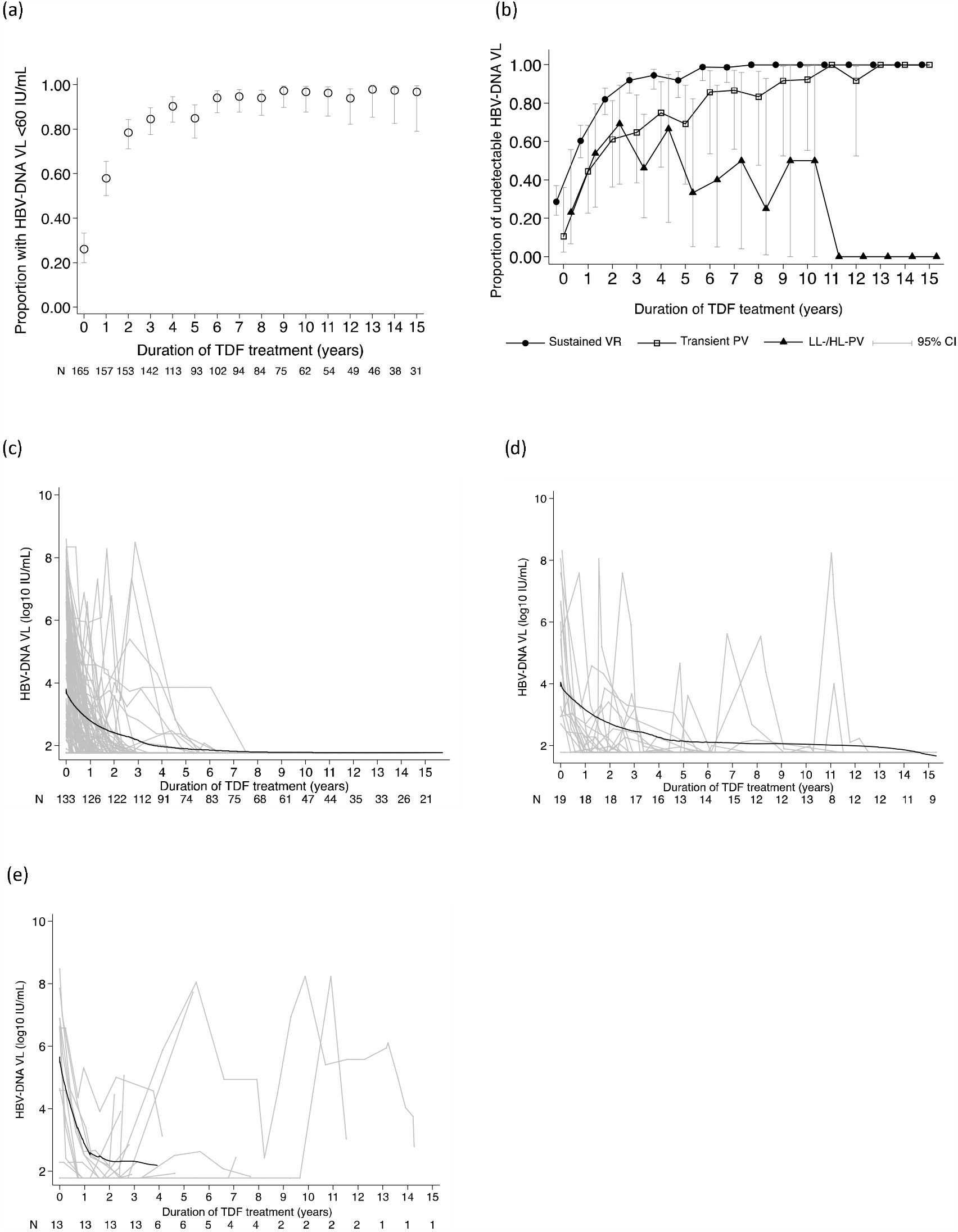
Viral suppression during tenofovir (TDF)-treatment in (a) all patients and (b) different profiles of HBV replication; and HBV-DNA viral loads during TDF-treatment in patients with (c) sustained virological response (VR), (d) transient persistent viremia (PV), and (e) low-level/high-level persistent viremia (LL-/HL-PV). Individual levels are expressed as gray lines.

163 (98.8%) patients were able to achieve virological response after a median 0.90 years (IQR=0.39-1.74) of follow-up and of them, 30 (18.2%) had at least one visit with detectable HBV-DNA after achieving virological response. The number of visits with detectable HBV-DNA for these patients were distributed as follows: 1, *n*=17; 2, *n*=7; 3, *n*=2; 4, *n*=2; 10, *n*=1; and, 18, *n*=1. Risk factors for viral persistence over time are given in Table 2. In multivariable analysis, positive HBeAg status at baseline (*p*=0.04), lower nadir CD4+ cell counts (*p*=0.002) and undetectable HIV-VL (*p*<0.001) were significantly associated with viral persistence.

**Table 2.**
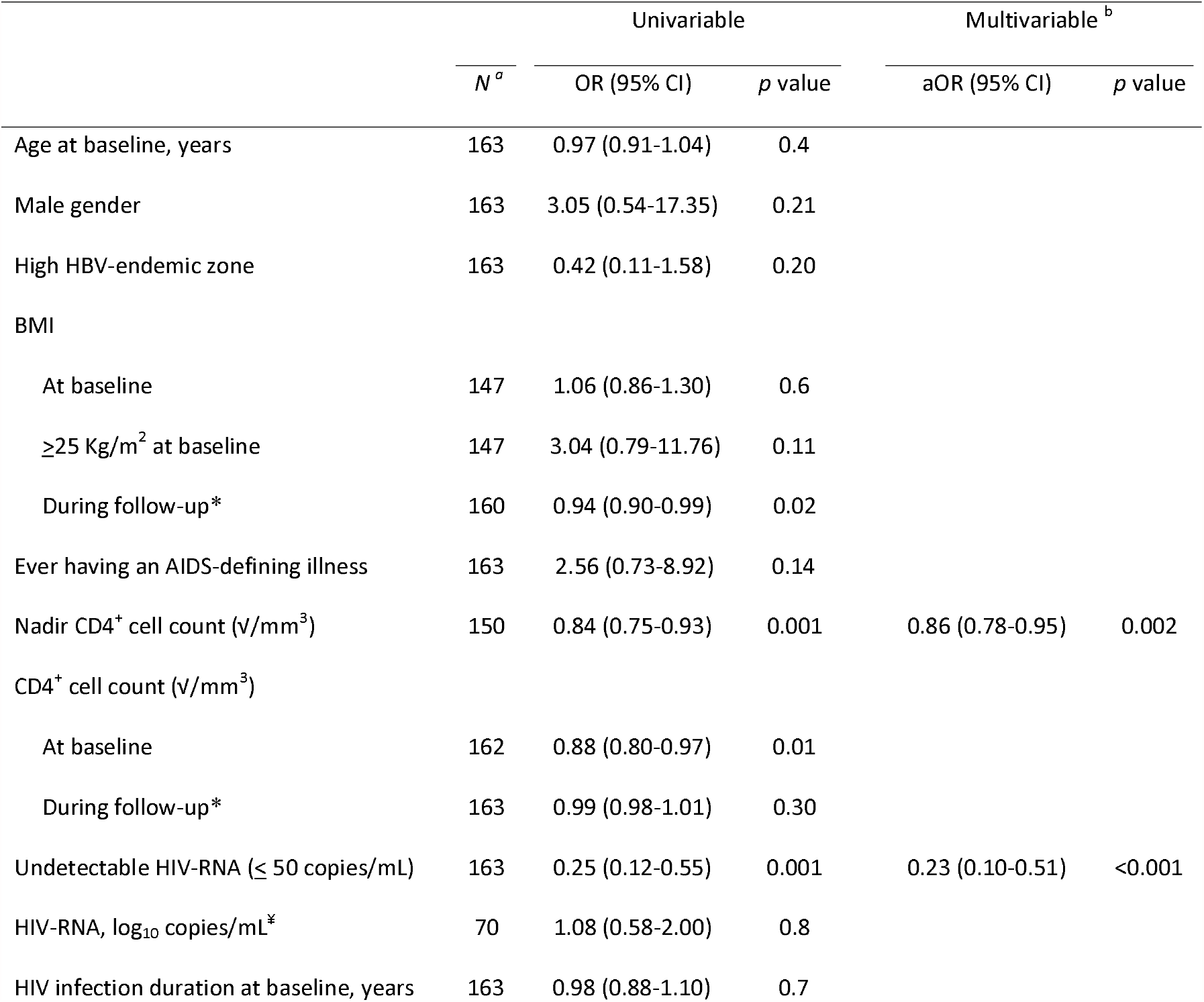
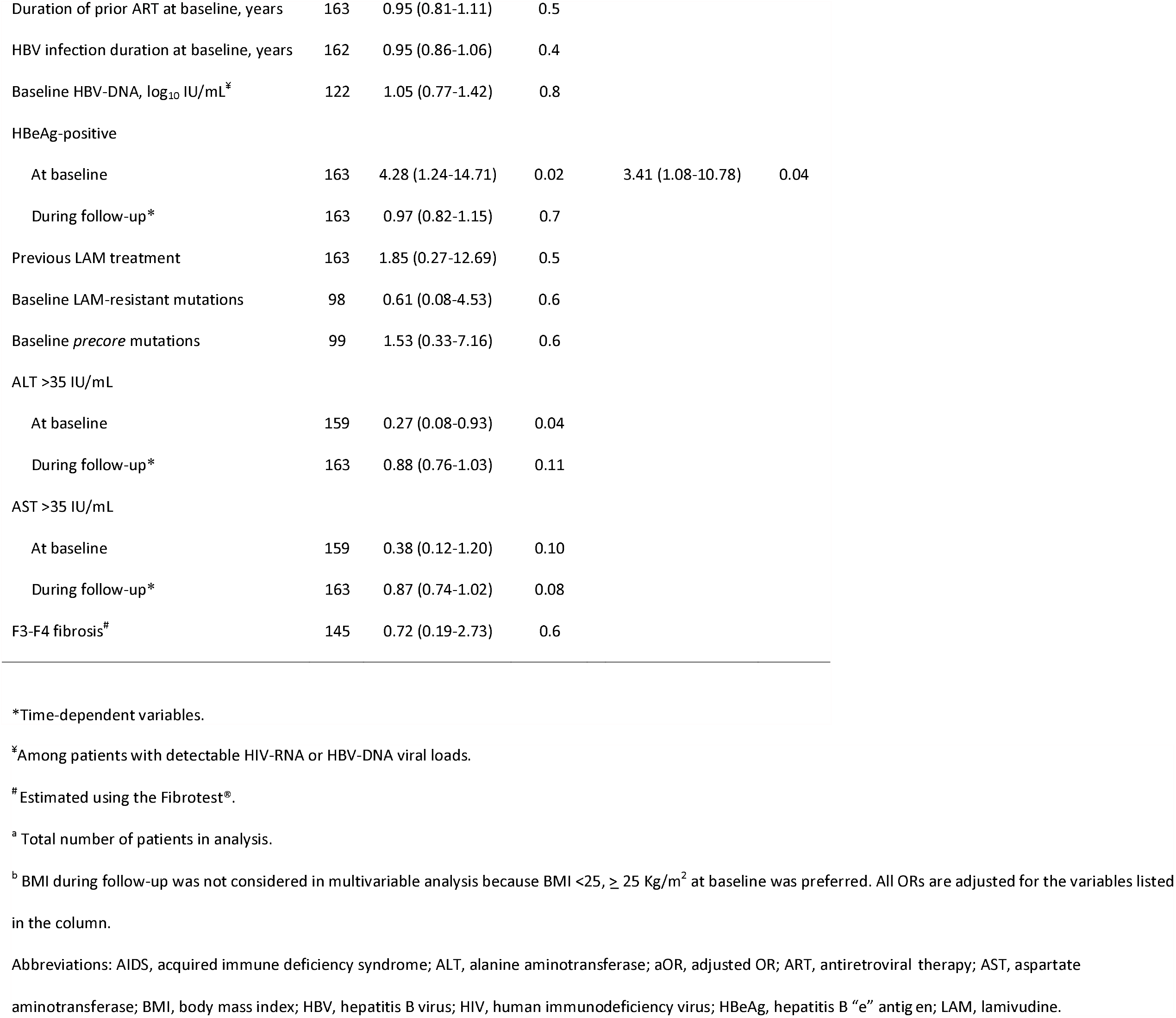
Determinants of viral persistence (≥60 IU/mL) after having achieved virological response (*N*=163).

We then characterized HBV replication profiles based on HBV-VLs at the end of follow-up. Of the 152 (92.1%) patients with undetectable HBV-VL, 133 (87.5%) had remained undetectable (sustained-VR) and 19 (12.5%) had become detectable (median peak HBV-VL=2.54 log_10_ IU/mL, range=1.79-8.04) after having achieved undetectable HBV-VL (transient-PV). Among those with transient-PV, 10 had more than one visit with detectable HBV-DNA, with the time from first to last detectable HBV DNA measurement lasting a median 1.0 year (range=0.12-7.0). Of the 13 (7.9%) patients with detectable HBV-VL at the end of the follow-up, 9 had LL-PV (median HBV-VL=272 IU/mL, range=67-1,341) and 4 had HL-PV (range HBV-VL=3.91-7.72 log_10_ IU/mL). Of these patients, 11 had achieved HBV-VL <60 at least once during follow-up (LL-PV, *n*=8; HL-PV, *n*=3). The proportion of patients with undetectable HBV-VLs are summarized between groups in Figure 1b, while changes in HBV-VLs are given for patients with sustained-VR in Figure 1c, transient-PV in Figure 1d and LL-/HL-PV in Figure 1e.

At baseline, patients with transient-PV profile, as compared to sustained-VR, were significantly more likely to be HBeAg-positive (p=0.04), to have detectable HIV-VL (p=0.04), lower nadir CD4+ cell count (p=0.002), higher BMI (p=0.01), and shorter cumulative LAM treatment prior to TDF-initiation (p=0.03) (Table 3). Patients with LL-/HL-PV profiles had significantly higher baseline ALT (p=0.007) and AST levels (p=0.03), when compared to those with either sustained-VR or transient-PV. During follow-up, patients with sustained-VR profile, compared to transient-PV, were significantly more likely to achieve virological response at 24 months of therapy (p=0.03), showed higher frequency of suppressed HIV-VL (p<0.001) and lower frequency of liver fibrosis progression from mild or moderate fibrosis (F0-F1-F2) to more advanced fibrosis (F3-F4; p=0.04) (Table 4). When including only individuals with two or more visits with detectable HBV-DNA (after VR) in the transient-PV group, similar differences between groups were observed (data not shown). Individuals with LL-/HL-PV profiles, when compared to all other profiles, were significantly more likely to have shorter duration of follow-up (p<0.001), higher 12-month (p<0.001) and 24-month change (p<0.001) in HBV-VL, higher maximum decrease (p<0.001) and increase (p<0.001) of ALT levels from baseline, and higher ALT levels at the last follow-up visit (p<0.001). Moreover, individuals with LL-/HL-PV profiles, compared to sustained-VR, had a lower frequency of undetectable HIV-VL for all visits during follow-up (Table 4).

**Table 3.**
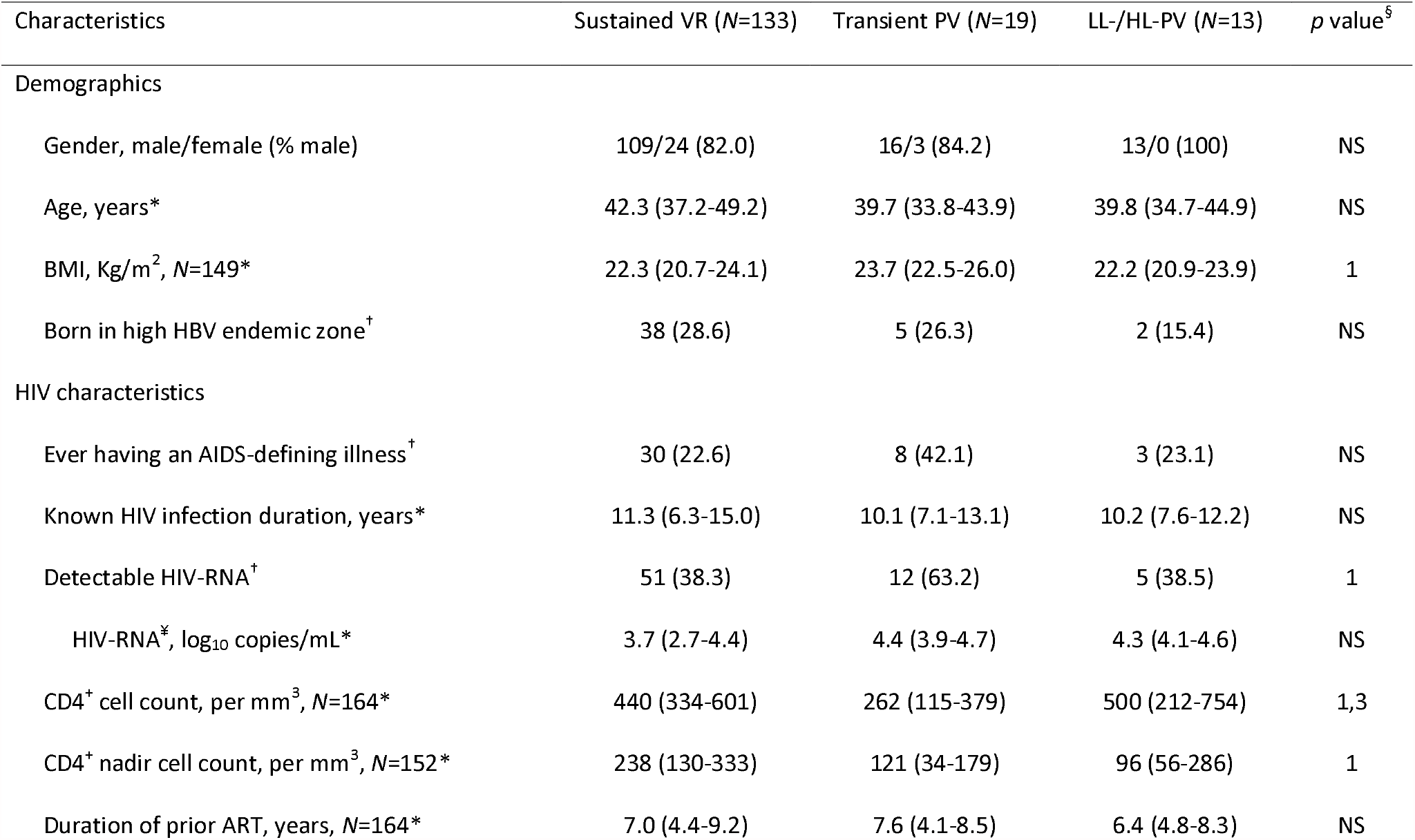
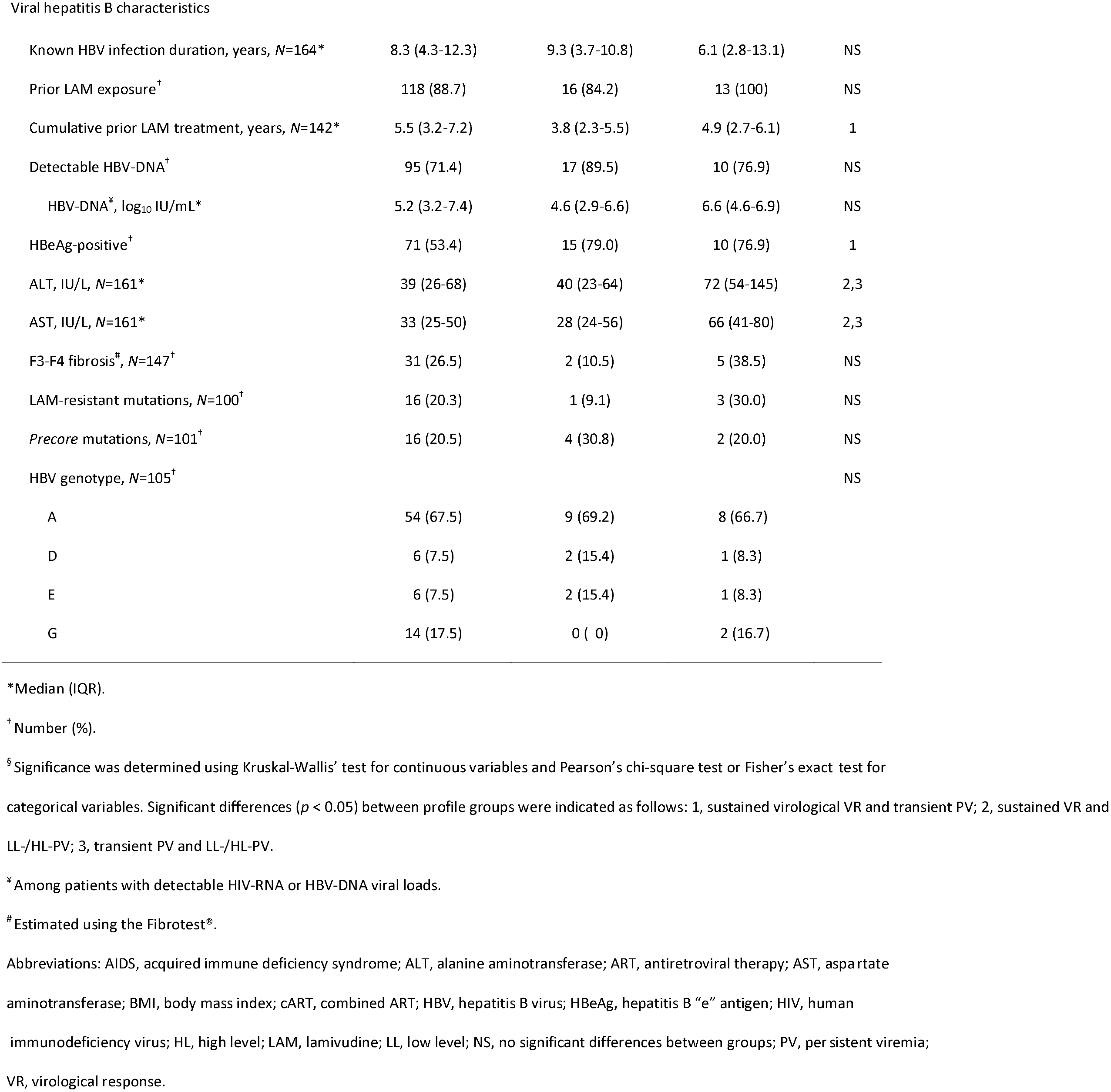
Baseline characteristics of hepatitis B virus replication profiles.

| Characteristics | Sustained VR (N=133) | Transient PV (N=19) | LL-/HL-PV (N=13) | *p* value<sup>§</sup> |
| --- | --- | --- | --- | --- |
| Demographics |  |  |  |  |
| Gender, male/female (% male) | 109/24 (82.0) | 16/3 (84.2) | 13/0 (100) | NS |
| Age, years* | 42.3 (37.2-49.2) | 39.7 (33.8-43.9) | 39.8 (34.7-44.9) | NS |
| BMI, Kg/m<sup>2</sup>, N=149* | 22.3 (20.7-24.1) | 23.7 (22.5-26.0) | 22.2 (20.9-23.9) | 1 |
| Born in high HBV endemic zone<sup>†</sup> | 38 (28.6) | 5 (26.3) | 2 (15.4) | NS |
| HIV characteristics |  |  |  |  |
| Ever having an AIDS-defining illness<sup>†</sup> | 30 (22.6) | 8 (42.1) | 3 (23.1) | NS |
| Known HIV infection duration, years* | 11.3 (6.3-15.0) | 10.1 (7.1-13.1) | 10.2 (7.6-12.2) | NS |
| Detectable HIV-RNA<sup>†</sup> | 51 (38.3) | 12 (63.2) | 5 (38.5) | 1 |
| HIV-RNA<sup>‡</sup>, log<sub>10</sub> copies/mL* | 3.7 (2.7-4.4) | 4.4 (3.9-4.7) | 4.3 (4.1-4.6) | NS |
| CD4<sup>+</sup> cell count, per mm<sup>3</sup>, N=164* | 440 (334-601) | 262 (115-379) | 500 (212-754) | 1,3 |
| CD4<sup>+</sup> nadir cell count, per mm<sup>3</sup>, N=152* | 238 (130-333) | 121 (34-179) | 96 (56-286) | 1 |
| Duration of prior ART, years, N=164* | 7.0 (4.4-9.2) | 7.6 (4.1-8.5) | 6.4 (4.8-8.3) | NS |
| Viral hepatitis B characteristics |  |  |  |  |
| Known HBV infection duration, years, *N*=164* | 8.3 (4.3-12.3) | 9.3 (3.7-10.8) | 6.1 (2.8-13.1) | NS |
| Prior LAM exposure<sup>†</sup> | 118 (88.7) | 16 (84.2) | 13 (100) | NS |
| Cumulative prior LAM treatment, years, *N*=142* | 5.5 (3.2-7.2) | 3.8 (2.3-5.5) | 4.9 (2.7-6.1) | 1 |
| Detectable HBV-DNA<sup>†</sup> | 95 (71.4) | 17 (89.5) | 10 (76.9) | NS |
| HBV-DNA<sup>‡</sup>, log<sub>10</sub> IU/mL* | 5.2 (3.2-7.4) | 4.6 (2.9-6.6) | 6.6 (4.6-6.9) | NS |
| HBeAg-positive<sup>†</sup> | 71 (53.4) | 15 (79.0) | 10 (76.9) | 1 |
| ALT, IU/L, *N*=161* | 39 (26-68) | 40 (23-64) | 72 (54-145) | 2,3 |
| AST, IU/L, *N*=161* | 33 (25-50) | 28 (24-56) | 66 (41-80) | 2,3 |
| F3-F4 fibrosis<sup>#</sup>, *N*=147<sup>†</sup> | 31 (26.5) | 2 (10.5) | 5 (38.5) | NS |
| LAM-resistant mutations, *N*=100<sup>†</sup> | 16 (20.3) | 1 (9.1) | 3 (30.0) | NS |
| *Precore* mutations, *N*=101<sup>†</sup> | 16 (20.5) | 4 (30.8) | 2 (20.0) | NS |
| HBV genotype, *N*=105<sup>†</sup> |  |  |  | NS |
| A | 54 (67.5) | 9 (69.2) | 8 (66.7) |  |
| D | 6 (7.5) | 2 (15.4) | 1 (8.3) |  |
| E | 6 (7.5) | 2 (15.4) | 1 (8.3) |  |
| G | 14 (17.5) | 0 ( 0) | 2 (16.7) |  |

\*Median (IQR).
<sup>†</sup> Number (%).
<sup>§</sup> Significance was determined using Kruskal-Wallis' test for continuous variables and Pearson's chi-square test or Fisher's exact test for categorical variables. Significant differences ( $p < 0.05$ ) between profile groups were indicated as follows: 1, sustained virological VR and transient PV; 2, sustained VR and LL-/HL-PV; 3, transient PV and LL-/HL-PV.
<sup>¥</sup> Among patients with detectable HIV-RNA or HBV-DNA viral loads.
<sup>#</sup> Estimated using the Fibrotest®.
Abbreviations: AIDS, acquired immune deficiency syndrome; ALT, alanine aminotransferase; ART, antiretroviral therapy; AST, aspartate aminotransferase; BMI, body mass index; cART, combined ART; HBV, hepatitis B virus; HBeAg, hepatitis B "e" antigen; HIV, human immunodeficiency virus; HL, high level; LAM, lamivudine; LL, low level; NS, no significant differences between groups; PV, persistent viremia; VR, virological response.

**Table 4.**
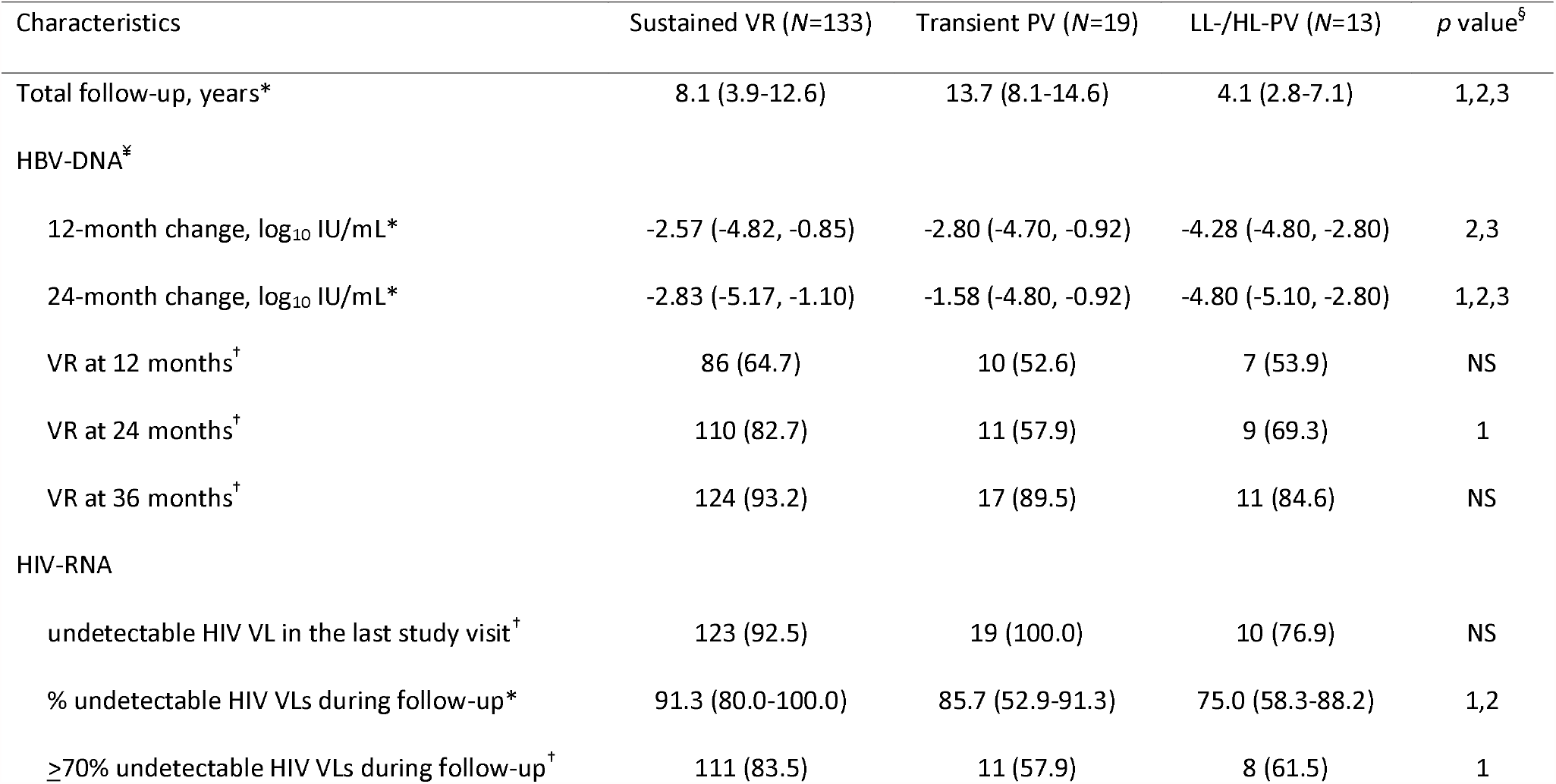
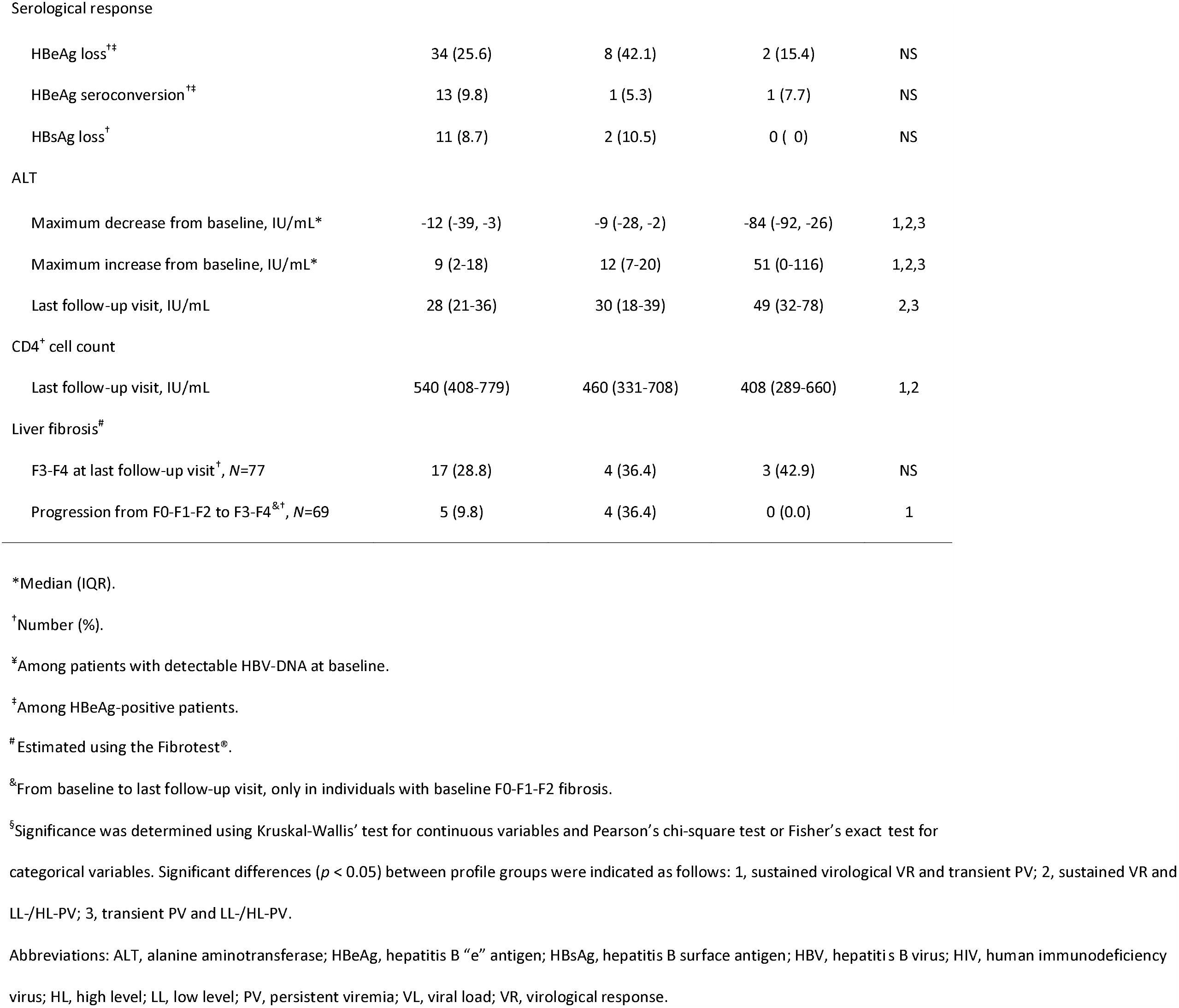
Clinical and serological characteristics of hepatitis B virus replication profiles during follow-up.

| Characteristics | Sustained VR (N=133) | Transient PV (N=19) | LL-/HL-PV (N=13) | *p* value<sup>§</sup> |
| --- | --- | --- | --- | --- |
| Total follow-up, years* | 8.1 (3.9-12.6) | 13.7 (8.1-14.6) | 4.1 (2.8-7.1) | 1,2,3 |
| HBV-DNA<sup>‡</sup> |  |  |  |  |
| 12-month change, log<sub>10</sub> IU/mL* | -2.57 (-4.82, -0.85) | -2.80 (-4.70, -0.92) | -4.28 (-4.80, -2.80) | 2,3 |
| 24-month change, log<sub>10</sub> IU/mL* | -2.83 (-5.17, -1.10) | -1.58 (-4.80, -0.92) | -4.80 (-5.10, -2.80) | 1,2,3 |
| VR at 12 months<sup>†</sup> | 86 (64.7) | 10 (52.6) | 7 (53.9) | NS |
| VR at 24 months<sup>†</sup> | 110 (82.7) | 11 (57.9) | 9 (69.3) | 1 |
| VR at 36 months<sup>†</sup> | 124 (93.2) | 17 (89.5) | 11 (84.6) | NS |
| HIV-RNA |  |  |  |  |
| undetectable HIV VL in the last study visit<sup>†</sup> | 123 (92.5) | 19 (100.0) | 10 (76.9) | NS |
| % undetectable HIV VLs during follow-up* | 91.3 (80.0-100.0) | 85.7 (52.9-91.3) | 75.0 (58.3-88.2) | 1,2 |
| ≥70% undetectable HIV VLs during follow-up<sup>†</sup> | 111 (83.5) | 11 (57.9) | 8 (61.5) | 1 |
| Serological response |  |  |  |  |
| HBeAg loss<sup>†‡</sup> | 34 (25.6) | 8 (42.1) | 2 (15.4) | NS |
| HBeAg seroconversion<sup>†‡</sup> | 13 (9.8) | 1 (5.3) | 1 (7.7) | NS |
| HBsAg loss<sup>†</sup> | 11 (8.7) | 2 (10.5) | 0 ( 0) | NS |
| ALT |  |  |  |  |
| Maximum decrease from baseline, IU/mL* | -12 (-39, -3) | -9 (-28, -2) | -84 (-92, -26) | 1,2,3 |
| Maximum increase from baseline, IU/mL* | 9 (2-18) | 12 (7-20) | 51 (0-116) | 1,2,3 |
| Last follow-up visit, IU/mL | 28 (21-36) | 30 (18-39) | 49 (32-78) | 2,3 |
| CD4<sup>+</sup> cell count |  |  |  |  |
| Last follow-up visit, IU/mL | 540 (408-779) | 460 (331-708) | 408 (289-660) | 1,2 |
| Liver fibrosis<sup>#</sup> |  |  |  |  |
| F3-F4 at last follow-up visit<sup>†</sup>, N=77 | 17 (28.8) | 4 (36.4) | 3 (42.9) | NS |
| Progression from F0-F1-F2 to F3-F4<sup>&†</sup>, N=69 | 5 (9.8) | 4 (36.4) | 0 (0.0) | 1 |

\*Median (IQR).
<sup>†</sup>Number (%).
<sup>‡</sup>Among patients with detectable HBV-DNA at baseline.
<sup>#</sup>Among HBeAg-positive patients.
<sup>#</sup> Estimated using the Fibrotest®.
<sup>&</sup>From baseline to last follow-up visit, only in individuals with baseline F0-F1-F2 fibrosis.
<sup>§</sup>Significance was determined using Kruskal-Wallis' test for continuous variables and Pearson's chi-square test or Fisher's exact test for categorical variables. Significant differences ( $p < 0.05$ ) between profile groups were indicated as follows: 1, sustained virological VR and transient PV; 2, sustained VR and LL-/HL-PV; 3, transient PV and LL-/HL-PV.
Abbreviations: ALT, alanine aminotransferase; HBeAg, hepatitis B "e" antigen; HBsAg, hepatitis B surface antigen; HBV, hepatitis B virus; HIV, human immunodeficiency virus; HL, high level; LL, low level; PV, persistent viremia; VL, viral load; VR, virological response.

### HBeAg- and HBsAg-seroclearance and its relation to viral persistence during TDF-containing ART

Of the 96 HBeAg-positive patients at study inclusion, 44 lost HBeAg (cumulative incidence: 45.8%; 95%CI=35.6%-56.3%) after a median 4.9 years (IQR=3.0-9.0) of follow-up (incidence rate=6.1/100 person-years). Of them, 2 reverted back to HBeAg-positive serology and 4 changed serostatus multiple times until ending follow-up with HBeAg-negative serology. Of the 42 patients ending follow-up with HBeAg-negative serology, acquisition of anti-HBe antibodies (anti-HBeAb) was achieved in 14 (33.3%) patients either at the same visit as HBeAg-seroclearance (n=10) or from 1.7-11.4 years after HBeAg-seroclearance (n=4). Of these patients, 6 (42.9%) lost anti-HBeAb by the end of follow-up.

No difference was observed in HBeAg-seroclearance between those who were able to achieve VR during follow-up versus those who never achieved VR (7.4 versus 3.7/100 person-years respectively, *p*=0.33, Figure 2a). Of those who had HBeAg-seroclearance, 34 belonged to the sustained-VR, 8 transient-PV profile, and 2 LL-PV groups.

**Figure 2.**
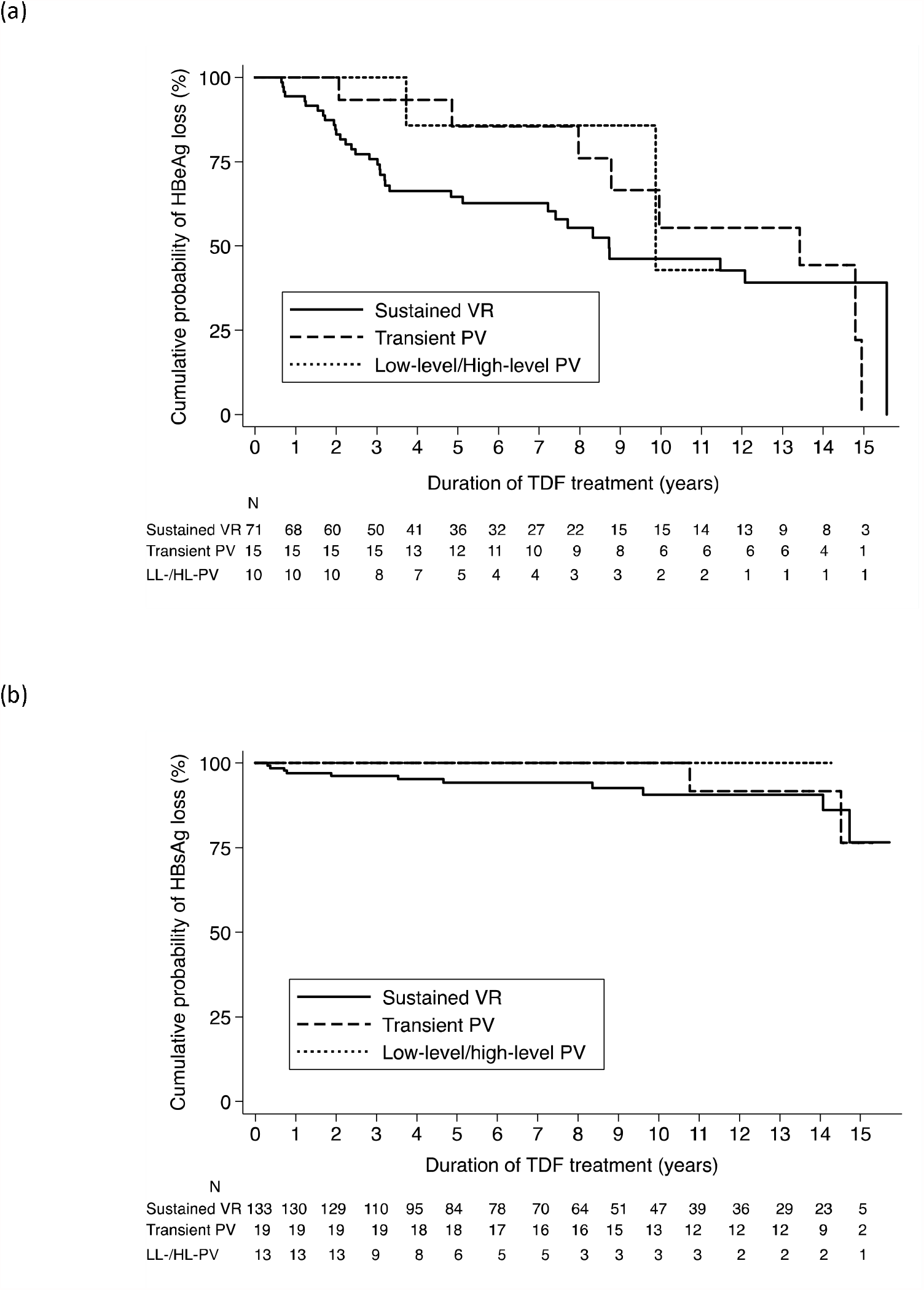
Cumulative probability of (a) hepatitis B ‘e’ antigen (HBeAg) and (b) hepatitis B surface antigen (HBsAg) seroclearance, according to different profiles of HBV replication.

A total of 13 patients lost HBsAg (cumulative incidence: 7.9%; 95%CI=4.3%-13.1%) after a median 7.5 years (IQR=3.8-11.2) of follow-up (incidence rate=0.9/100 person-years). Of these patients, two reverted back to HBsAg-positive serology and one changed serostatus multiple times until ending follow-up with HBsAg-positive serology. Of the 10 patients ending follow-up with HBsAg-negative serology, acquisition of anti-HBsAb was achieved in six (60.0%) either at the same visit as HBsAg-seroclearance (n=3) or from 0.9-6.7 years after HBsAg-seroclearance (n=3). Additionally, 6 patients who lost HBsAg were HBeAg-positive at baseline. For these patients, median time from HBeAg-seroclearance to HBsAg-seroclearance was 4.8 years (range=0.5-13.5).

HBsAg-seroclearance was only observed in those who achieved VR, yet this rate was not significantly different from those who were never able to achieve VR (1.0 versus 0/100 person-years, respectively, *p*=0.49, Figure 2b). Of those who had HBsAg-seroclearance, 11 belonged to the sustained-VR and 2 transient-PV profile groups.

### Severe liver-related morbidity and mortality and its relation to viral persistence during TDF-containing ART

At TDF initiation, 4 patients had already experienced a severe liver-related event [portal hypertension n=1; HCC, n=2; haemorrhagic necrosis of liver, n=1]. Of them, 2 had sustained VR, 1 had transient PV and 1 had HL-PV. During TDF treatment, 3 patients developed portal hypertension (all with sustained VR profiles), 1 hepatorenal syndrome (HL-PV profile), 3 HCC (2 with sustained VR and 1 with LL-PV profiles), and 2 haemorrhagic necrosis of liver (1 with sustained VR and 1 with transient PV). Two deaths were the result of HCC (1 with sustained VR and 1 with LL-PV profile) and one from decompensated liver with HCC and complications due to septic shock (sustained-VR profile). All three patients who developed HCC during follow-up had METAVIR F3-F4 fibrosis at TDF initiation, as measured by the FibroTest®. Ultrasound revealed the presence of hepatic nodules for all three patients, along with steatosis (n=2) and portal vein thrombosis (n=1).

## Discussion

Using longitudinal data from one of the longest studies to date on TDF-use in HIV-HBV coinfected individuals, we observed that viral persistence continues to occur throughout TDF-treatment, with 18.2% of patients exhibiting detectable HBV-DNA after having achieved virological response. Nevertheless, the probability of having detectable HBV-DNA decreased as the duration of TDF increased and ≤6% of patients consistently had detectable HBV-DNA every year after 6 years of TDF. These data support the durability of HBV suppression associated with TDF-use and suggest that viral persistence does not mitigate long-term viral suppression.

The extent of persistent viremia in individuals with chronic HBV infection varies considerably across studies. When defining persistence based on HBV-DNA levels at a maximum 24 months of follow-up, we found that only 8% of patients presented with LL-/HL-PV. If minimum follow-up is extended to 60 months, where most HIV-HBV coinfected individuals appeared to have achieved HBV virological suppression, only 4% of patients had LL-PV and no patient had HL-PV. The proportion of patients with HBV persistence in our study is considerably lower when compared to other prospective studies in HIV-HBV coinfected, reporting 14% to 54%.^8,9,12^ However, these studies used inability to achieve virological response at 12-months as the basis for persistent viremia, which, given our data and others,^9^ is too short a timeframe to define persistence. When defining persistence based on any detection of HBV-VL after achieving VR, we found that 20% of individuals had at one point persistent viremia. In the GS-US-174-0102 and GS-US-174-0103 registration studies in HBV mono-infected patients, this type of persistence occurred very rarely (0.9%).^23^

We observed that median CD4^+^ cell count at last follow-up visit and the frequency of undetectable HIV-VL over time were significantly higher in patients with sustained-VR compared to other profiles, while patients who had higher CD4^+^ counts for longer periods of time were able to more frequently suppress HBV-DNA replication after achieving initial virological response. Indeed, immunosuppression as an underlying factor for viral persistence has been evoked in other studies.^12,15^ Previous research has suggested that HIV-HBV coinfected patients have lower levels of HBV-specific CD4^+^ T-cell responses,^24,25^ intrahepatic T cells, Kupfer cells and NK cells,^26^ as well as reduced intrahepatic inflammatory activity when compared to HIV-positive patients without HBV infection. Taken together, these data highlight the importance of immunoregulation to control HBV replication and could explain why persistence occurs more frequently in HIV-HBV coinfected versus HBV mono-infected patients.

Detectable HIV-RNA viremia was also more often observed during treatment for individuals with transient-PV, LL- and HL-PV profiles and was strongly associated with HBV viral persistence after achieving virological response. Similar findings have been observed by others.^9,12^ In general, detectable HIV-RNA, as determined by most commercial assays, is either due to developing HIV resistant variants or inadequate adherence to ART.^27^ Given that no consistent mutation pattern for TDF resistance has yet to be observed,^4^ detectable HBV-DNA would likely be the result of poor adherence.^28^ Much of the concordance of simultaneously detectable HIV-RNA and HBV-DNA would then likely be the result of insufficient adherence. We did not collect data on HIV resistance and our data on HBV sequences are limited to the first 8 years of follow-up, hence we are unable to confirm this speculation. TDF plasma concentrations were indeed lower or even undetectable for individuals with profiles of LL/HL-PV in our previous study,^8^ while others have found lower concentrations of intracellular drug levels of tenofovir-diphosphate, a measure of long-term adherence, for HIV-HBV coinfected individuals with HBV persistent viremia during TDF-treatment.^29^ Nevertheless, considering that HIV-RNA was undetectable for 71% of visits when HBV-VL was detectable (after initial HBV virological response), adherence alone cannot fully explain HBV persistent viremia.

For individuals with persistent viremia, it is clear that the vast majority are able to eventually suppress HBV-DNA. It remains debatable to what extent persistent viremia leads to major long-term serological consequences. We observed that individuals with LL- and HL-PV profiles were never able to achieve HBsAg-seroclearance. The reasons for this finding are not entirely clear. Large rebounds in HBV-DNA replication have been shown to be associated with HBsAg-seroclearance, but is usually accompanied with ALT flares and occurs mostly in HBV treatment cessation studies or during the course of natural infection;^30,31^ increases in ALT levels were rarely observed during HBV viral persistence in our study population of TDF-treated individuals. It could be that ability to achieve sustained VR is a proxy for tighter control of HBV viral activity in general; however, further evidence would be needed to confirm this finding. Nevertheless, it should be noted that only 7.9% of the entire study population were able to exhibit HBsAg-seroclearance, even after up to 15 years of follow-up. The lack of function cure would appear to be more a general problem in treated coinfected patients.

The clinical consequences of persistent HBV viremia are also fairly unknown. From several studies during the natural history of chronic HBV infection, consistently high levels of HBV-DNA (i.e. ≥10,000 copies/mL) during later phases of HBV infection are mostly associated with developing HCC and cirrhosis.^32^ Expectedly, our data showed a low incidence of clinical liver-related outcomes, with no strong evidence that PV *per se* is associated with higher rates of liver-related morbidity and mortality.

This study has several strengths, including possibly the longest follow-up to date in either HIV-HBV coinfected^8,9,13,18^ and HBV-monoinfected patients^14,33^ with consistently measured virological and serological markers of HBV. However, certain limitations need to be addressed. First, the time-dependent definitions of HBV replications profiles might be inadequate for under 3 years of follow-up. Therefore, we decided to also use persistent viremia after initial VR as a complementary definition of persistence. Second, we did not measure plasma TDF concentrations to assess adherence, so we cannot determine if those patients with viral persistence were adherent to ART or whether a decrease in frequency of persistence was due to improved adherence over time. Third, our data represent a population that is highly ART-experienced and more immunosuppressed compared to contemporary patient populations, but still actively seen in outpatient settings. Finally, since genotypic data were not collected at later years of TDF use in our cohort, we are unable to determine whether viral persistence is related to certain mutational patterns.^34^ From our last analysis, there were only 9 additional patients exhibiting an HBV-VL >1000 IU/mL, which would be ideal for sequencing, and the small numbers would unlikely reveal any other mutation patterns explaining viral persistence during TDF use.

In conclusion, we demonstrate that TDF is able to suppress HBV-DNA to undetectable levels in the majority of ART-experienced HIV-HBV coinfected patients undergoing up to 15 years of TDF-containing ART. Nevertheless, a low proportion of patients do exhibit HBV viral persistence. A small proportion of patients achieved HBV functional cure, which was exclusively observed in those with sustained HBV-VLs. This low percentage of serological responders is quite similar to HBV mono-infected patients undergoing prolonged TDF treatment^14^ and highlights the need to identify novel HBV treatment strategies against HBV. Still, evaluating the relationship between HBV persistent viremia and more severe clinical outcomes is warranted in larger, longitudinal cohort studies.

## Data Availability

A full list of data citations are available by contacting the corresponding author.

## Acknowledgements

The authors are grateful to the patients and the clinical teams for their commitment to the French HIV-HBV Cohort. This study was sponsored by the Institut de Médecine et d’Epidémiologie Appliquée (IMEA). L.N.C.D. was awarded a post-doctoral fellowship from the France REcherche Nord&sud Sida-hiv Hépatites (ANRS).

## Role of each author

L.N.C.D. was responsible for the statistical analysis, interpretation of the data, and drafting the manuscript. S.M., A.G. and C.D. were responsible for interpretation of the data and drafting the manuscript. H.R., P.M., C. L-C., and J.C. acquired data for the cohort, assisted in interpreting data, and gave critical revisions of the manuscript. K.L. helped design, conceptualize, and obtain funding for the French HIV-HBV cohort study, coordinated data collection, and drafted parts of the manuscript. A.B. coordinated data analysis, gave important comments on data interpretation, drafted the manuscript, and provided critical revisions of the manuscript. All authors approved the final version.

## Transparency declarations

None to declare.

## Supplementary Data

Figure S1 is available as Supplementary data.

