## Supplementary figures and images for "Persistent HBV replication and serological response during up to fifteen years of tenofovir-based antiretroviral therapy in HIV-hepatitis B coinfected patients: a multicenter prospective cohort study"

### Supplemental figure S1

**Supplementary Figure S1. Patient flow**

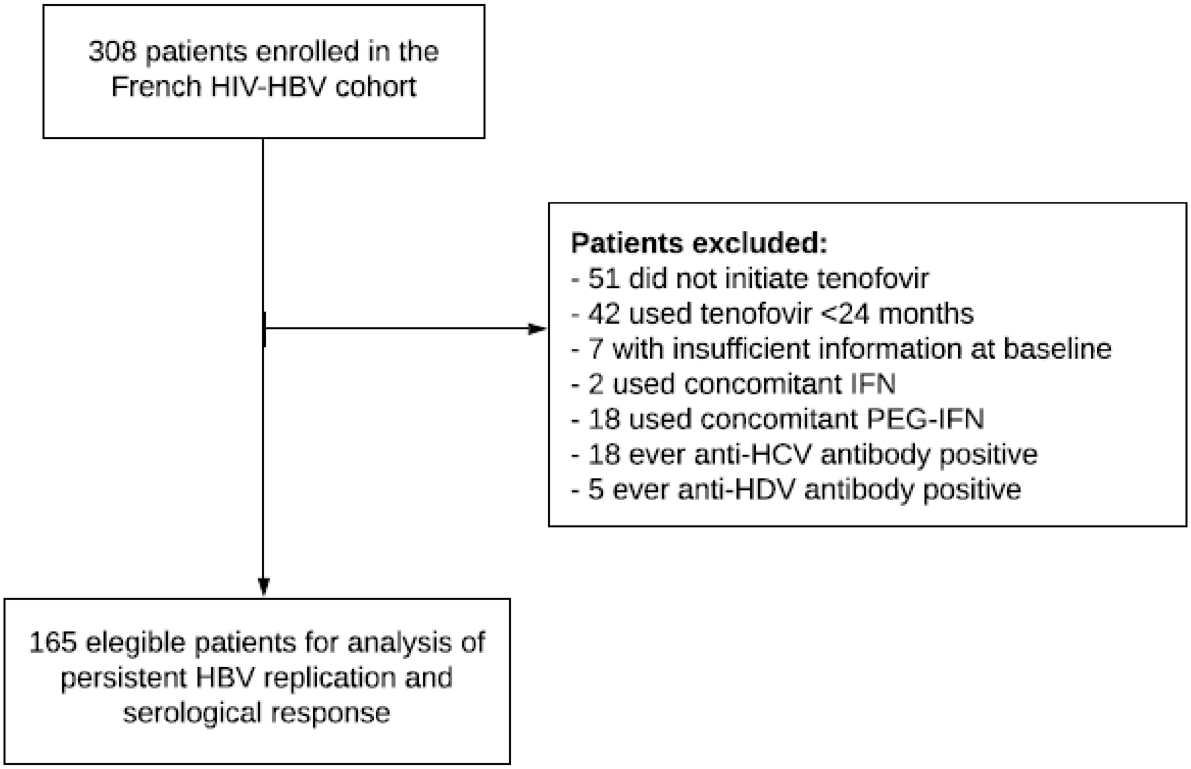
